# Heavy load carrying and musculoskeletal health: An exploratory study of sand miners in Pokhara, Kaski District, Nepal

**DOI:** 10.1101/2020.11.11.20227652

**Authors:** Aybüke Koyuncu, Michael N. Bates, Ziva Petrin, Myles Cope, Sandra I. McCoy, Ndola Prata, Tula Ram Sijali, Carisa Harris-Adamson

## Abstract

**Background:** Trends in urbanization contribute to the growing global demand for raw construction materials. The health effects of load carrying among occupational groups that mine and carry sand and stone used for construction of roads and buildings remains poorly understood.

**Methods:** We conducted an exploratory cross-sectional study among a convenience sample of sand miners working at an excavation site on the Seti River in Pokhara, Nepal. Load carrying weight, duration, and frequency were used to categorize miners as having “low” or “high” load-carrying exposures. Probable musculoskeletal disorders (MSDs) were identified using self-reported symptoms of moderate to severe musculoskeletal pain, as well as physical examinations.

**Results:** The average loads carried by female and male sand miners weighed 66.3 kg and 87.3 kg, respectively. Among all participants (N=42), 45.2% reported moderate to severe musculoskeletal pain in at least one body region and 16 (38.1%) had probable MSDs identified using specified case criteria. The prevalence of MSDs was lower among miners carrying, on average, heavier loads compared to those carrying lighter loads (OR_a_=0.29; 95% CI: 0.05, 1.8), possibly indicative of the healthy worker survival effect. Miners carrying loads for longer durations and frequencies had higher odds of MSDs compared with those carrying for shorter durations and frequencies.

**Conclusion:** Despite the pervasiveness of load carrying as an income generating activity throughout the developing world, these populations remain largely excluded from global occupational health agendas. Larger epidemiologic studies are needed to justify action to protect the health and safety of these unrecognized and understudied groups.

## 1. INTRODUCTION

Rapid population growth combined with increasing global urbanization contributes to the enormous demand for raw materials, such as sand and gravel, for use in concrete (1,2). By 2050, it is anticipated that there will be a need for at least 25 million kilometers of new roads, with 90% of all road construction expected to take place in developing countries (1). In order to meet the growing need for raw building materials in global development, occupations such as sand mining have become increasingly important. Developing countries, in which the anticipated demand for raw building materials is highest, are least likely to have mechanization to facilitate the extraction and transport of sand and stone deposits from the environment (1).

Although a number of studies have reported associations of heavy load carrying with self-reported pain and/or degenerative changes in the cervical spine, the majority of existing research has examined load carriage on top of the head (3). Additionally, loads carried in Sub-Saharan Africa, where most previous such research has been conducted, are typically 25-35 kg, significantly less than the average loads carried by sand miners in Nepal, where our study is based (3). To our knowledge, no studies have examined the potential detrimental musculoskeletal impacts of sand mining work practices.

We aimed to address the existing knowledge gap on the health impacts of heavy load carrying by collecting descriptive and quantitative data on heavy load carrying practices and their association with adverse musculoskeletal health among sand miners in Nepal. For this purpose, we conducted an exploratory cross-sectional study among men and women working as sand miners at a site along the Seti River in Pokhara, Nepal.

## 2. METHODS

### 2.1 Setting

During each Monsoon season in Nepal (generally, May to September), heavy rains cause rivers to flood, washing sand and rocks of various sizes down from the Annapurna mountain range of the Himalayas. One of these rivers, the Seti, travels at high velocity through narrow gorges, with the flow rate subsiding where the river gorge opens out into a much wider area within the boundaries of the city of Pokhara. The reduced flow rate causes the sand and rocks to sink, forming a high floodplain.

When the Monsoon rains subside, the floodplain site turns into a commercial sand mining operation (Figure I).

**Figure I.**
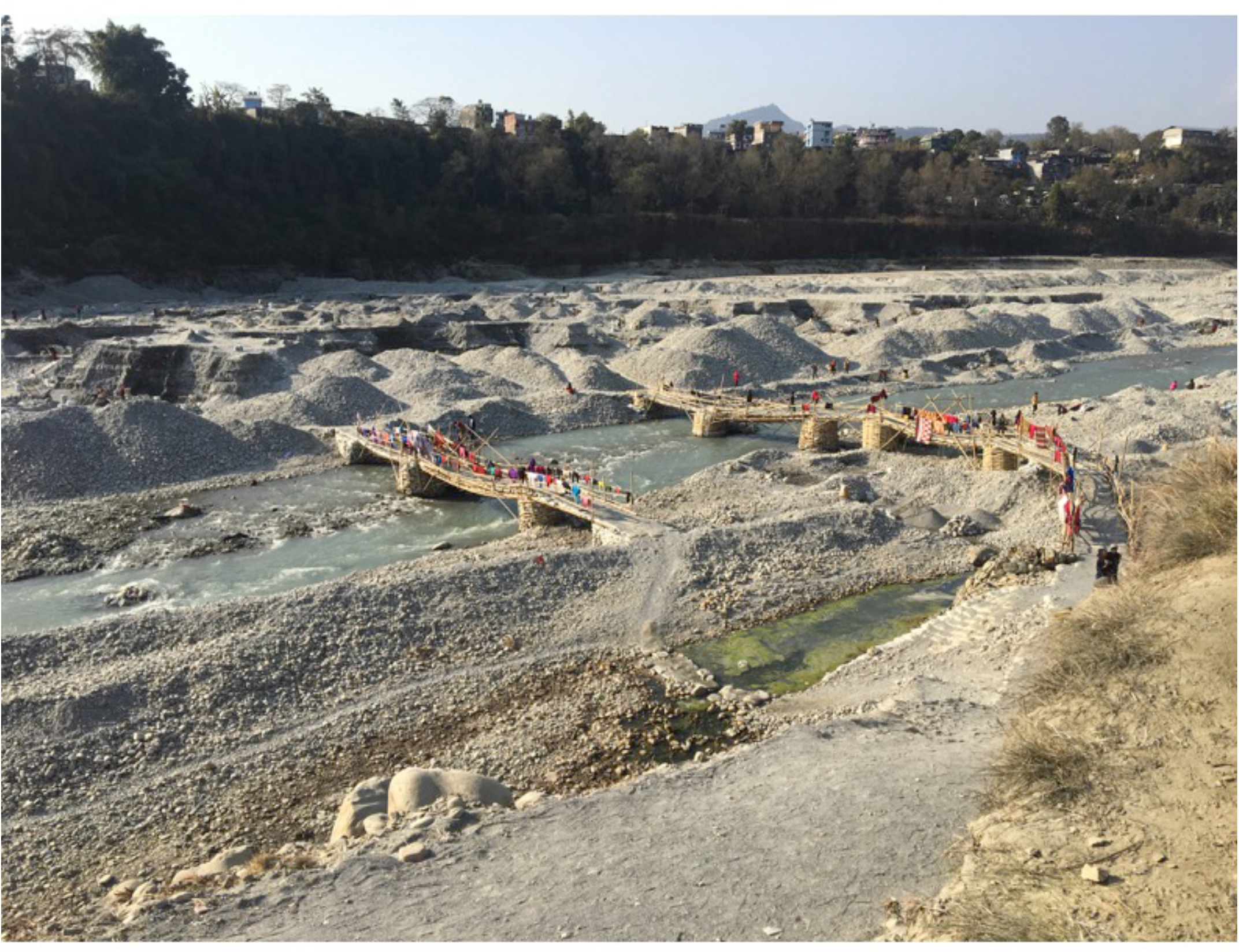
A sand mining site at the Seti River in Pokhara, Kaski district, Nepal.

Sand miners, both men and women, stake out “claims” and manually load and transport the sand and rocks across the river bed and up the river bank, where the loads are piled according to size. Men and women miners transport these loads on their backs using a cone-shaped basket called a “doko”, supported by a circular band called a “namlo” looped around the forehead.

Although dokos and namlos are a typical way of transporting loads in Nepal, given the frequency and weight of loads carried by sand miners, this occupational group represents an extreme example of such carriage. Workers load the excavated material into trucks, which transport it for use in the construction of roads and buildings. Miners usually operate in small groups and are paid according to the number of truck-loads they fill.

Miner groups operate with division of tasks often differentiated by sex. At designated mining sites along the river, women use sieves and other tools to manually sort and sift through sand and stones. Women also frequently carry loads short distances within their excavation area to create piles of sand and stone of similar size that are ready to be transported to collection trucks. Both men and women transport the materials across the riverbed and up the bank, and there is variation in the weights of loads and distances travelled. Both groups use namlos and dokos to transport their loads, but those performing the task of sorting often travel shorter distances and more frequently. The starting point for a load is when the doko (basket) is placed on an approximately waist-high mound filled with the material to be transported. The miner steps with his or her back to the mound and, using the namlo, hoists the basket onto their back. 2.2 Study Population

Men and women over the age of 18 working at a sand mining site along the Seti River in Pokhara were eligible for recruitment. A convenience sample of men and women was selected among those observed to be carrying a load of sand at the mining site. Due to the unwillingness of sand mining site managers to allow research staff to actively recruit study participants at the mining site, recruitment of study participants relied heavily on word of mouth.

### 2.3 Data Collection

Data collection took place during December 2017-January 2018. Eligible study participants who provided informed consent completed a survey, administered in Nepali by trained local interviewers. The survey covered a range of topics, including socio-demographics, relationship status, load-carrying activities, musculoskeletal and reproductive health, and, among women, childbearing history. The height and weight of each participant, as well as the weight of the load being carried at time of study recruitment, were obtained using measuring tape and scales.

Participants were asked whether they experienced symptoms of musculoskeletal pain, defined as pain lasting 3 or more days in the neck, head, back, knees, feet and/or ankles, in the last year. Individuals reporting pain were asked about pain frequency (number of days in the last month) and its severity on a 0-10 numeric pain scale (NPS) in each affected body region. Participants who reported pain in the last year, with 3 or more days of pain in the last month, rated as a severity 4 or greater on the rating scale, were asked to undergo a non-invasive examination by a licensed Nepali physiotherapist for evaluation of the presence of musculoskeletal abnormalities in the areas of reported pain. Physical examinations were conducted according to a pre-specified study protocol, either on the day of the survey or an agreed-upon date in the weeks following survey completion. The physical examination included a variety of maneuvers to detect indicators of adverse musculoskeletal health, such as resisted agonist muscle contractions, localized pain or tenderness, or limited range of motion (ROM). Full examination details can be found in the Supplementary Materials.

### 2.4 Study Indicators

#### 2.4.1 Exposure: Heavy load carrying practices

Individual load carrying practices were quantified in terms of the weight of loads carried at the time of study recruitment, self-reported average load-carrying duration (number of minutes per load carried) and frequency (number of loads carried per week). A cumulative measure of load-carrying exposures in the last week was constructed by multiplying the load weight by duration and frequency. Each individual load-carrying exposure (weight, duration, frequency, cumulative load carrying) was dichotomized at the median exposure level to indicate “low” or “high” exposures.

#### 2.4.2 Outcome: Probable Work-Related Musculoskeletal Disorders

Self-reported pain and physical examination findings were used to classify participants for the possible presence of musculoskeletal disorders (MSDs) in three main body areas: neck, low back, and lower extremities. Additionally, based on prior studies showing a high incidence of spondylosis in groups with head-supported carriage (4–6), MSDs in the neck and low back groups were further divided into two groups: axial (neck or low back pain, which is likely to originate from local muscle, tendon, or bone) and radicular (neck or low back pain which radiates to an extremity, and may originate from compression of a spinal nerve root). Validated physical examination tests were used to create a set of classification criteria to identify probable work-related MSDs (Table I).

**Table I.** Case Criteria for Identification of Probable Work-Related Musculoskeletal Disorders (MSDs)

| Probable MSD | Self-Reported History | Diagnostic Criteria |
| --- | --- | --- |
| Nonspecific neck pain | Neck pain $\geq 4$ on NPS for $\geq 3$ days in last month | Negative Spurling test; and<br>Negative axial load test; and<br>Asymmetrical range of motion (ROM) or<br>tenderness to palpation in cervical paraspinals or upper trapezius |
| Cervicalgia with possible radiculopathy | Neck pain $\geq 4$ on NPS for $\geq 3$ days in last month | 1) Positive Spurling test (26,27); or<br>2) Positive axial load test (28); or<br>3) Asymmetrical reflex in the upper extremity; or |
| | | 4) Asymmetrical myotome/Manual muscle testing grade in the upper extremity; or<br>5) Myotome grade $\leq 3/5$ on manual muscle testing |
| Nonspecific low back pain | Low back pain $\geq 4$ on NPS for $\geq 3$ days in last month | Negative straight leg test; and<br>Negative Slump test; and<br>1) Positive SI maneuver testing; or<br>2) Asymmetrical reflex in the lower extremity; or<br>3) Asymmetrical myotome/Manual muscle testing grade in the lower extremity; or<br>4) Myotome grade $\leq 3/5$ on manual muscle testing in lower extremity<br>5) Asymmetrical ROM (side-bend or rotation) or tenderness to palpation in lumbar spine |
| Dorsalgia with radiculopathy | Low back pain $\geq 4$ on NPS for $\geq 3$ days in last month | 1) Positive Sciatica history; or<br>2) Positive straight leg (29,30) or<br>3) Positive Slump test;(30,31) or<br>4) Positive axial load test<br>AND<br>1) Asymmetrical reflex in the lower extremity; or<br>2) Asymmetrical myotome/Manual muscle testing grade in the lower extremity; or<br>3) Myotome grade $\leq 3/5$ on manual muscle testing in lower extremity |
| Knee MSD | Knee pain $\geq 4$ on NPS for $\geq 3$ days in last month | Tenderness to palpation in the: ITB, biceps femoris, patellar tendon, joint line/meniscus, medial collateral ligament, lateral collateral ligament<br>AND<br>1) Positive genu valgum/valgus; or<br>2) Abnormality in knee testing maneuvers: extensor lag, patellar mobility, patellar tracking with quadricep activation, MCL/LCL/ACL/PCL integrity, bounce home test, flexion with over pressure, McMurray Test, clicking, Apleys Compression Test |
| Ankle MSD | Foot/ankle pain $\geq 4$ on NPS for $\geq 3$ days in last month | 1) Positive windlass test (32); or<br>2) Positive single leg heel raise; or<br>3) Positive navicular test; or<br>4) Positive talar tilt test (33) |

There was no follow-up of participants and no data were collected on the outcomes of any subsequent medical consultations.

### 2.5 Statistical Analyses

Bivariate analyses between probable work-related MSDs, load carrying exposures and potential confounders of interest were conducted using chi-squared tests, Fisher’s exact tests, and t-tests, as appropriate. Following bivariate analyses, separate unadjusted and adjusted logistic regression models were utilized to examine the association between load carrying exposures and probable MSDs in at least one body region. Fully adjusted models, including potential confounders of the association between MSDs and load carrying contained age as a continuous variable and sex. Due to our limited sample size, other potential confounders, such as body mass index (BMI) and years worked as a sand miner, were not included in adjusted models. The positive predictive value and corresponding 95% confidence interval (CI) of self-reported history of pain for MSDs identified by physical exams were calculated by dividing the number of true positives (participants who were identified by physical examination as having a probable MSD) by the total number of true and false positives (participants reporting pain). To explore sex-based differences in the associations between load carrying exposures and probable MSDs, all unadjusted analyses were repeated in sub-groups defined by sex. Adjusted analyses were not repeated in sub-groups defined by sex due to our limited sample size.

In sensitivity analyses, three measures of load carrying practices (weight, duration, and frequency) were combined into a summary index of load carrying using principal component analysis (PCA). Data patterns between the various simultaneously occurring dimensions of load carrying were captured using the first component of the PCA. The summary index of load-carrying exposures was categorized as a binary variable indicating “low” or “high” load-carrying exposures split at the median score for the first component of the PCA. All analyses were conducted using STATA 15 (College Station, Texas).

### 2.6 Human Subjects Protection

The protocol for this study was approved by the Committee for the Protection of Human Subjects at the University of California, Berkeley, and the Nepal Health Research Council. No subject participated in this study before providing informed consent.

## 3. RESULTS

After the exclusion of 8 participants for whom data on their exposure, outcome, or covariates of interest was missing, the study population consisted of 20 females (mean age 35.3 (SD=7.4)) and 22 males (mean age 39.3 (SD=12.1)). Demographics of participants are shown in Table II.

Participants reported working as sand miners for an average of 7.4 years (SD=7.8), averaging 6.3 days (SD=1.0) of such work in the last week and 10.8 hours work per work day (SD= 2.5). There was heterogeneity in body mass index (BMI) by sex, with 9% of men and 45% of women being overweight (≥25 kg/m^2^).

**Table II.**
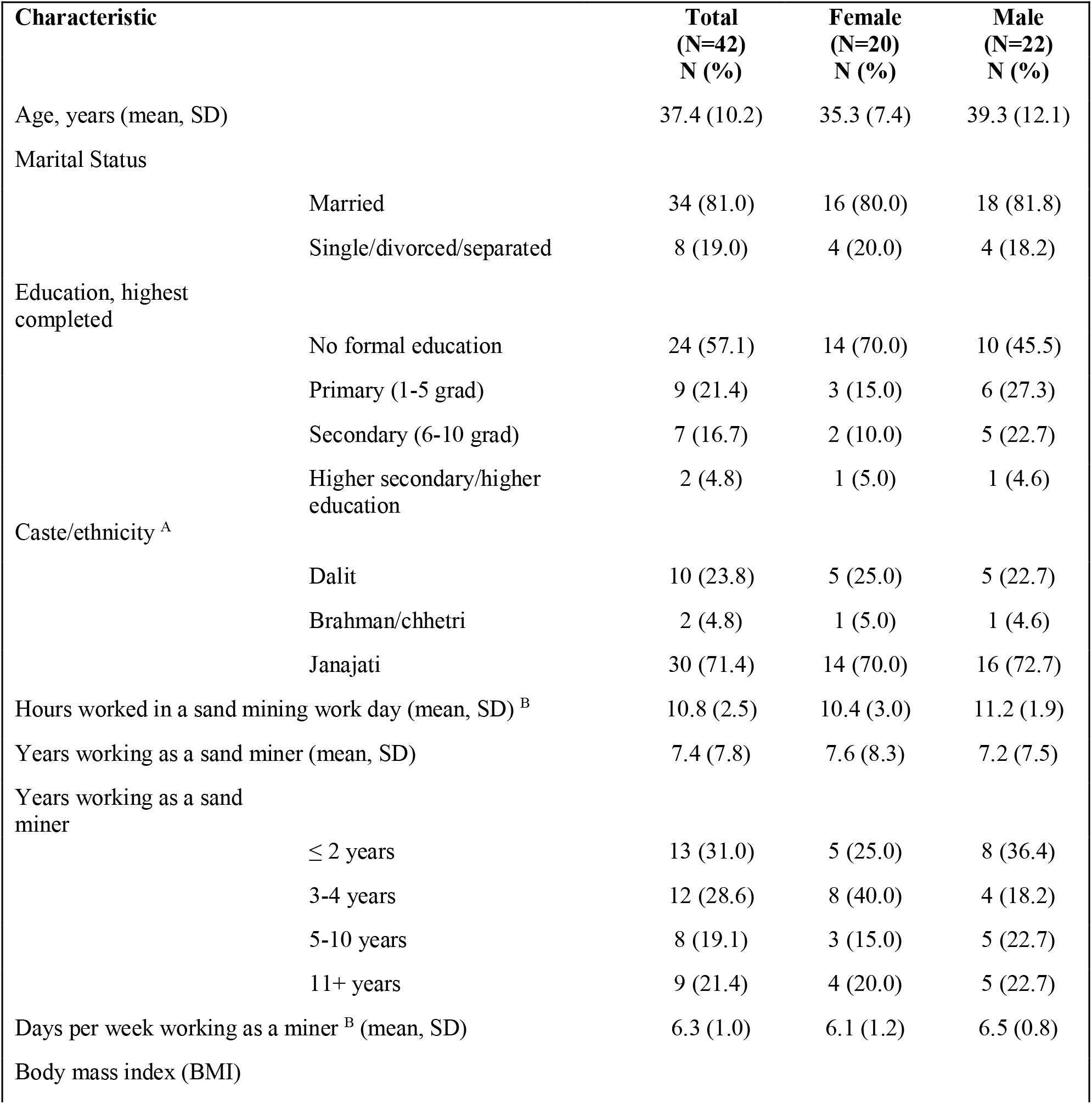
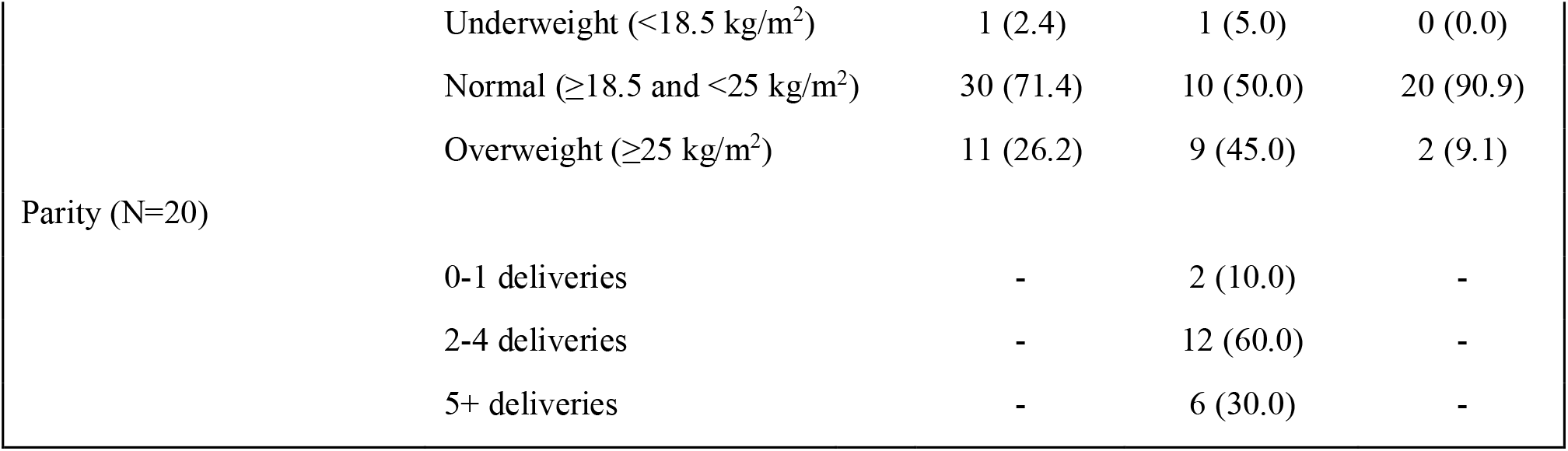
Sociodemographic characteristics of participants, Pokhara, Nepal, 2018. ^A^ Castes originate from the Hindu religion and are hierarchical groups based on social stratification. In Nepali culture, the Dalit represent a low caste group, Brahman/Chhetri are high caste Hindus, and Janjati are indigenous populations ^B^ Number of days in the last week worked as a sand miner

### 3.1 Load carrying exposures

Load-carrying statistics of the study population are summarized in Table III and Figure III.

**Table III.** Summary of load-carrying exposures among sand miners, Pokhara, Nepal, 2018.

| Load-carrying exposures | Mean (SD) |  |  |
| --- | --- | --- | --- |
|  | Overall (n=42) | Female (n=20) | Male (n=22) |
| Weight of load (kg) | 77.3<br>(17.4) | 66.3<br>(10.3) | 87.4<br>(16.4) |
| Weight of load (kg) to body mass<br>(kg) ratio | 1.4<br>(0.34) | 1.2<br>(0.32) | 1.6<br>(0.30) |
| Duration per load (mins) | 10.3<br>(6.0) | 9.3<br>(6.3) | 11.3<br>(5.6) |
| Frequency of loads per week | 304.4<br>(392.3) | 337.9<br>(565.2) | 274.0<br>(97.0) |
| Cumulative load-carrying index<br>(kg*mins/week) | 215,260.6<br>(213,910.2) | 178,111.4<br>(275,912.5) | 249,032.6<br>(133,975.8) |

**Figure III.**
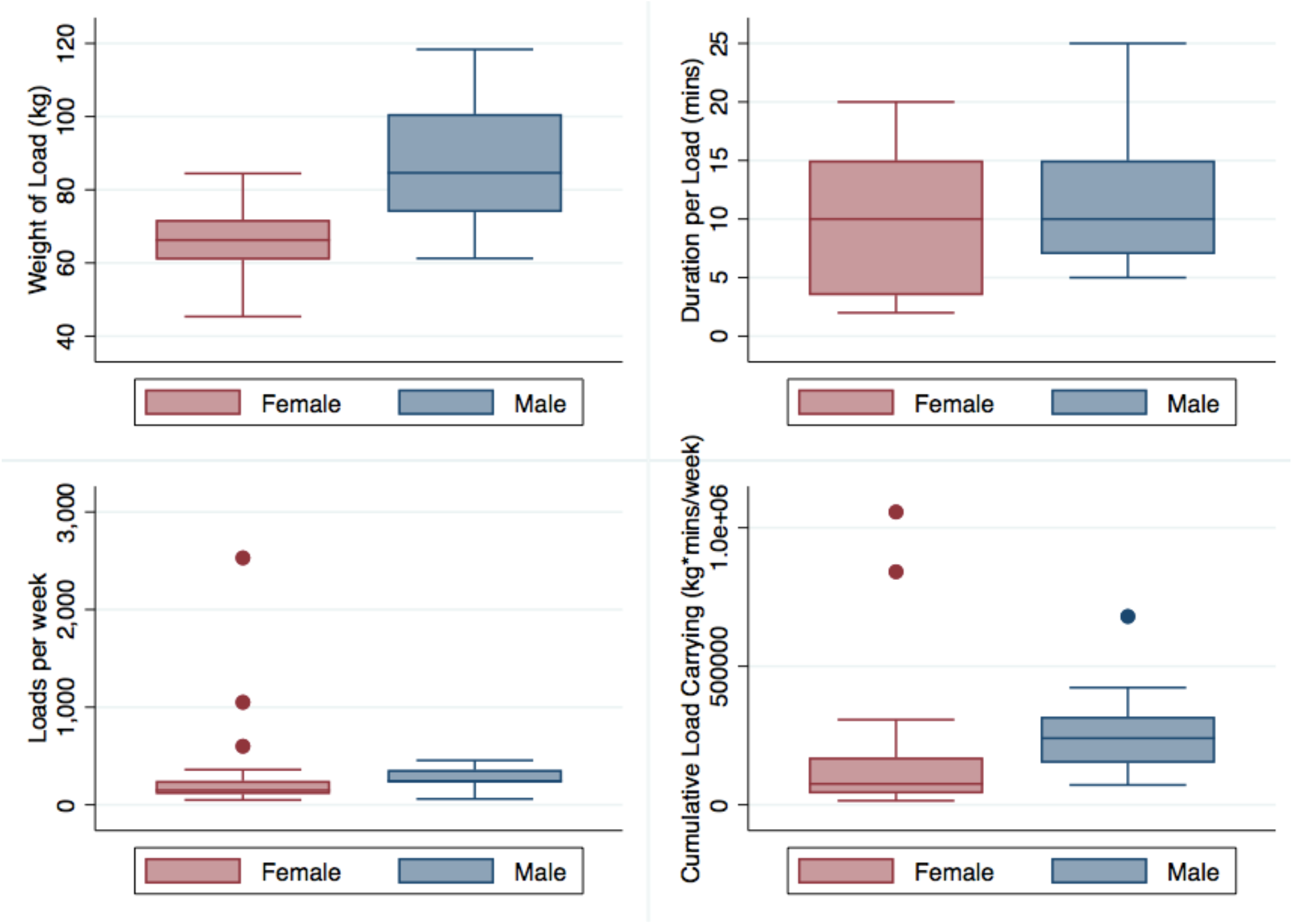
Load-carrying exposures among sand miners, Pokhara, Nepal, 2018.

The average loads carried by female and male sand miners weighed 66.3 kg (SD=10.3) and 87.4 kg (SD=16.4), respectively. Women carried, on average, 1.2 times their body weight (SD=0.32) and carried loads ranging from 45.4 to 84.5 kilograms. Men carried on average 1.6 times their body weight (SD=0.30), with loads ranging from 61.3 kilograms to 118.4 kilograms.

Compared to males, female sand miners carried on average lighter loads for shorter durations and higher frequencies. All study participants used namlos and dokos to carry loads and were carrying loads of sand or stones at the time of study recruitment.

### 3.2 Musculoskeletal health

In the overall study population, 19 participants (45.2%) triggered physical examinations based on self-reported musculoskeletal pain (≥ 4 on NPS for ≥ 3 days in last month) and, of those, 16 participants (38.1%) were assessed by physical examination as having a probable work-related MSD in at least one body region. Table IV shows the frequencies of self-reported pain and diagnosed MSDs.

**Table IV.**
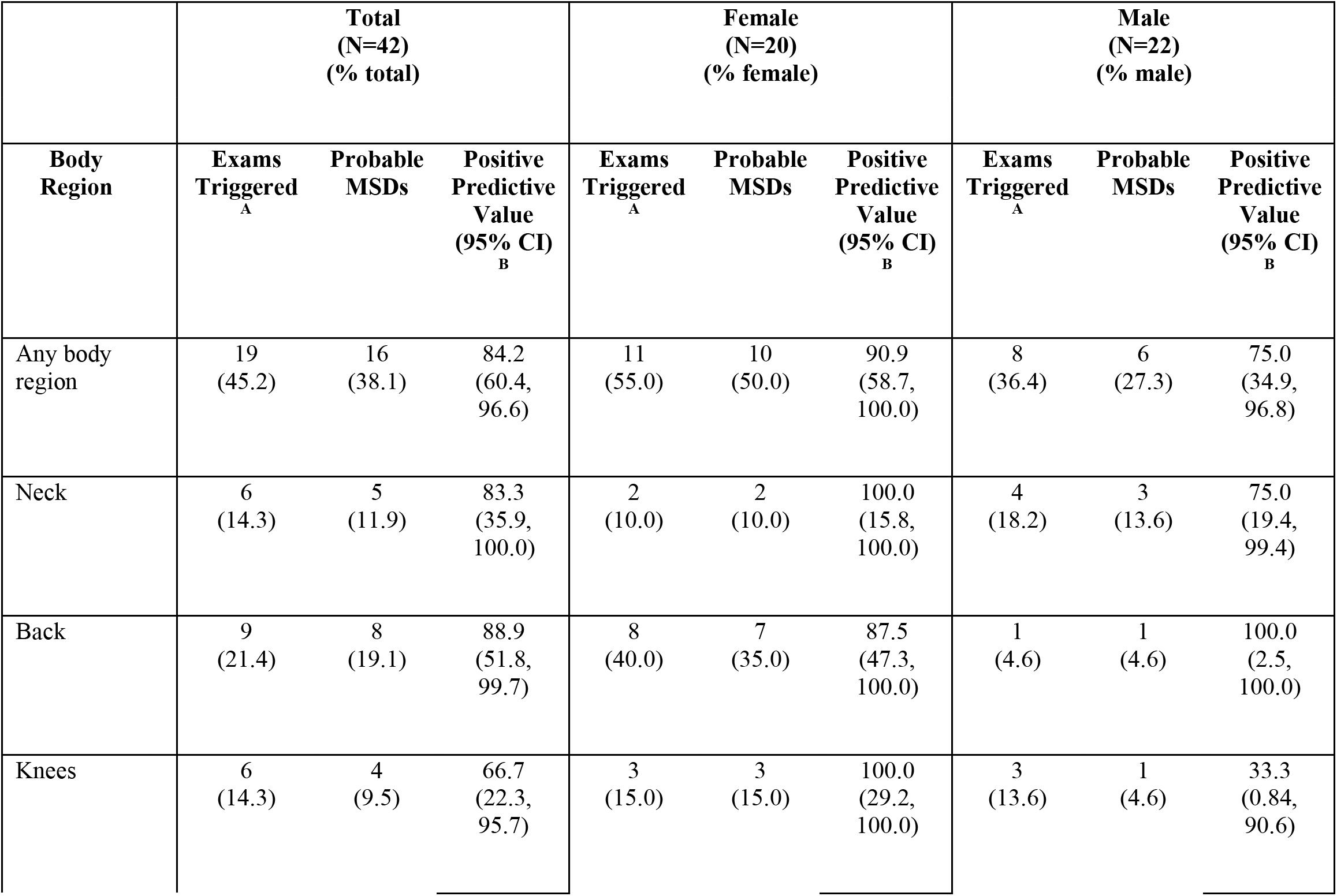
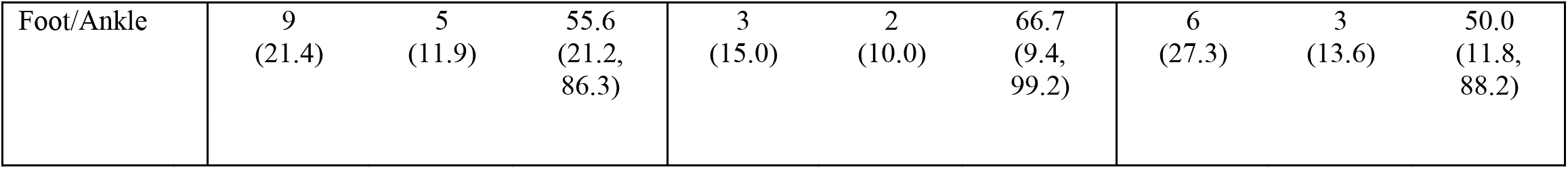
Summary of musculoskeletal pain and probable musculoskeletal disorders, Pokhara, Nepal, 2018. MSD: Musculoskeletal disorder CI: Confidence Interval ^A^ Self-reported Pain ≥ 4 on numeric pain scale (NPS) for ≥3 days in last month ^B^ One-sided 97.5% Confidence Interval, as appropriate

MSDs in the back (19.1% of participants) were the most prevalent, followed by the foot/ankle (11.9%) and neck (11.9%). We identified 4 participants with pain and exam findings suggestive of radicular involvement in the back and 2 in the neck region. Because of small numbers, axial and radicular MSD subgroups were combined in all further analyses.

Among the 19 sand miners who reported moderate to severe pain (≥ 4 on NPS for ≥ 3 days in last month), 84.2% had probable work-related MSDs in at least one body region (95% CI: 60.4, 96.6). The positive predictive value (PPV) of self-reported history of pain for identifying MSDs in the overall study population was highest for pain reported in the back (88.9%; 95% CI: 51.8, 99.7) and lowest for pain reported in the feet/ankle (55.6%; 95% CI: 21.2, 86.3). The feet/ankles had the lowest PPV among women (66.7%; 95% CI: 9.4, 99.2) while the knees had the lowest PPV among men (33.3; 95% CI: 11.8, 88.2). Self-reported pain was more likely to be indicative of probable work-related MSD among women compared to men in all body regions except the back.

Table V shows bivariate associations of MSD occurrence with load carrying and the potential confounders.

Females had a higher prevalence than males of MSDs in the back (35.0% vs 4.6%; p=0.02) (Table IV). Participants with MSDs in at least one body region had a higher average BMI compared to participants with no MSDs (24.9 kg/m^2^ vs 22.4 kg/m^2^; p=0.01). Older age was predictive of having an MSD in at least one body region (41.5 years vs 34.9 years; p=0.04), as well as in the knees/feet/ankles specifically (44.5 years vs 35.7 years; p=0.03).

### 3.3 Load carrying and musculoskeletal health

Odds of probable MSDs for high relative to low load-carrying groups varied depending on specific load-carrying exposure (Table VI).

**Table V.** Bivariate associations between musculoskeletal disorders, load carrying and potential confounders, Pokhara, Nepal, 2018 *p<0.10, **p<0.05 MSD: Musculoskeletal disorder BMI: Body mass index ^A^ (kg*mins/week). Low and high split based on median cumulative load carrying ^B^ P-values reported for chi-squared tests, Fisher’s exact tests, and t-tests, as appropriate

|  |  |  |  |  |
| --- | --- | --- | --- | --- |
|  | Heavy load carrying | Sex | Age, years | BMI (kg/m<sup>2</sup>) |

Table V. Bivariate associations between musculoskeletal disorders, load carrying and potential confounders, Pokhara, Nepal, 2018
|  |  | Low<br>cumulative<br>load<br>carrying <sup>A</sup><br>N (col %) | High<br>cumulative<br>load<br>carrying <sup>A</sup><br>N (col %) | p-<br>value<br><sub>B</sub> | Female<br>N (col<br>%) | Male<br>N (col<br>%) | p-<br>value<br><sub>B</sub> | Mean<br>(SD) | p-<br>value<br><sub>B</sub> | Mean<br>(SD) | p-<br>value<br><sub>B</sub> |
| --- | --- | --- | --- | --- | --- | --- | --- | --- | --- | --- | --- |
| **Any**<br>**body**<br>**region** | Probable<br>MSD<br>No | 13<br>(61.9) | 13<br>(61.9) | 1.0 | 10<br>(50.0) | 16<br>(72.7) | 0.13 | 34.9<br>(9.9) | 0.04** | 22.4<br>(2.7) | 0.01** |
|  | Yes | 8<br>(38.1) | 8<br>(38.1) |  | 10<br>(50.0) | 6<br>(27.3) |  | 41.5<br>(9.6) |  | 24.9<br>(3.0) |  |
| **Neck** | Probable<br>MSD<br>No | 19<br>(90.5) | 18<br>(85.7) | 0.63 | 18<br>(90.0) | 19<br>(86.4) | 1.00 | 36.5<br>(9.8) | 0.14 | 23.3<br>(3.0) | 0.75 |
|  | Yes | 2<br>(9.5) | 3<br>(14.3) |  | 2<br>(10.0) | 3<br>(13.6) |  | 43.8<br>(12.1) |  | 23.8<br>(1.4) |  |
| **Back** | Probable<br>MSD<br>No | 18<br>(85.7) | 16<br>(76.2) | 0.43 | 13<br>(65.0) | 21<br>(95.5) | 0.02** | 37.4<br>(10.6) | 0.96 | 23.0<br>(3.0) | 0.18 |
|  | Yes | 3<br>(14.3) | 5<br>(23.8) |  | 7<br>(35.0) | 1<br>(4.6) |  | 37.3<br>(9.1) |  | 24.7<br>(3.0) |  |
| **Knees,**<br>**Feet,**<br>**Ankles** | Probable<br>MSD<br>No | 16<br>(76.2) | 18<br>(85.7) | 0.43 | 16<br>(80.0) | 18<br>(81.8) | 1.00 | 35.7<br>(10.3) | 0.03** | 23.0<br>(3.0) | 0.14 |
|  | Yes | 5<br>(23.8) | 3<br>(14.3) |  | 4<br>(20.0) | 4<br>(18.2) |  | 44.5<br>(6.0) |  | 24.8<br>(3.1) |  |

In unadjusted analyses, the average weight of the load had a protective association with odds of MSDs (OR=0.18; 95% CI: 0.04, 0.71). The association between load weight and odds of MSDs was slightly attenuated in adjusted analyses (OR_a_=0.29; 95% CI: 0.05, 1.8). In contrast, higher load-carrying duration and frequency corresponded to higher odds of probable MSDs in adjusted analyses, although confidence intervals included the null. Among both men and women, fewer cases of MSDs were identified among miners carrying heavier loads compared to those carrying lighter loads (Table VII).

In unadjusted and adjusted sensitivity analyses exploring a summary index of load carrying generated using PCA, miners in the higher load carrying group had equal or lower odds of MSDs compared to those in the lower load carrying group (Table S1).

**Table S1.** Association between a summary index of load-carrying and probable work-related MSDs, Pokhara, Nepal, 2018 *p<0.10, **p<0.05 MSD: Musculoskeletal disorder; OR: Odds ratio ^A^ Summary measure of load-carrying generated using principle component analysis of load weight, duration, and frequency ^B^ Adjusted for age and sex

| Body region | Load Carrying <sup>A</sup> | Cases of Probable MSDs<br>N (col %) | Non-Cases<br>N (col %) | Unadjusted OR<br>(95% CI) | Adjusted <sup>B</sup> OR<br>(95% CI) |
| --- | --- | --- | --- | --- | --- |
| Any body region | Low | 9<br>(56.3) | 12<br>(46.2) | 1.00 (ref) | 1.00 (ref) |
|  | High | 7<br>(43.8) | 14<br>(53.9) | 0.67<br>(0.19, 2.3) | 0.70<br>(0.17, 2.9) |

**Table VI.** Association between load-carrying exposures and probable work-related musculoskeletal disorders, Pokhara, Nepal, 2018 *p<0.10, **p<0.05 MSD: Musculoskeletal disorder; OR: Odds ratio ^A^ Adjusted for age and sex

|  |  | Cases of Probable MSDs<br>N (col %) | Non-Cases<br>N (col %) | Unadjusted OR<br>(95% CI) | Adjusted<sup>A</sup> OR<br>(95% CI) |
| --- | --- | --- | --- | --- | --- |
| **Average weight of load (kg)** | Low | 12<br>(75.0) | 9<br>(34.6) | 1.00 (ref) | 1.00 (ref) |
|  | High | 4<br>(25.0) | 17<br>(65.4) | 0.18**<br>(0.04, 0.71) | 0.29<br>(0.05, 1.8) |
| **Average load carry duration (mins)** | Low | 6<br>(37.5) | 11<br>(42.3) | 1.00 (ref) | 1.00 (ref) |
|  | High | 10<br>(62.5) | 15<br>(57.7) | 1.2<br>(0.34, 4.4) | 1.4<br>(0.3, 6.1) |
| **Average # of loads/Week** | Low | 8<br>(50.0) | 12<br>(46.2) | 1.00 (ref) | 1.00 (ref) |
|  | High | 8<br>(50.0) | 14<br>(53.9) | 0.86<br>(0.25, 3.0) | 1.9<br>(0.4, 9.6) |
| **Cumulative load carrying (kg\*mins/week)** | Low | 13<br>(50.0) | 8<br>(50.0) | 1.00 (ref) | 1.00 (ref) |
|  | High | 13<br>(50.0) | 8<br>(50.0) | 1.0<br>(0.29, 3.5) | 1.1<br>(0.24, 5.5) |

**Table VII.** Association between individual load carrying exposures and probable work-related MSDs in subgroups defined by sex, Pokhara, Nepal, 2018 *p<0.10, **p<0.05 MSD: Musculoskeletal disorder; OR: Odds ratio

|  |  | Females |  |  | Males |  |  |
| --- | --- | --- | --- | --- | --- | --- | --- |
|  |  | Cases of Probable MSDs<br>N (row %) | Non-Cases<br>N (row %) | Unadjusted OR<br>(95% CI) | Cases of Probable MSDs<br>N (row %) | Non-Cases<br>N (row %) | Unadjusted OR<br>(95% CI) |
| **Average weight of load (kg)** | Low | 7<br>(70.0) | 3<br>(30.0) | 1.00 (ref) | 6<br>(54.6) | 5<br>(45.5) | 1.00 (ref) |
|  | High | 3<br>(30.0) | 7<br>(70.0) | 0.18*<br>(0.03, 1.2) | 0<br>(0.0) | 11<br>(100.0) | NA |
| **Average load carry duration (mins)** | Low | 4<br>(44.4) | 5<br>(55.6) | 1.00 (ref) | 2<br>(25.0) | 6<br>(75.0) | 1.00 (ref) |
|  | High | 6<br>(54.6) | 5<br>(45.5) | 1.5<br>(0.26, 8.8) | 4<br>(28.6) | 10<br>(71.4) | 1.2<br>(0.17, 8.7) |
| **Average # of loads/Week** | Low | 3<br>(30.0) | 7<br>(70.0) | 1.00 (ref) | 2<br>(20.0) | 8<br>(80.0) | 1.00 (ref) |
|  | High | 7<br>(70.0) | 3<br>(30.0) | 5.4*<br>(0.80, 36.9) | 4<br>(33.3) | 8<br>(66.7) | 2.0<br>(0.28, 14.2) |

## 4. DISCUSSION

In, to our knowledge, the first study to collect occupational health data from sand miners, we quantified the workloads, as well as the prevalence of musculoskeletal morbidity experienced by miners in Pokhara, Nepal. Our data demonstrate the immense workloads undertaken by sand miners and provide preliminary information on associations of these workloads with MSDs.

Given the culturally distinctive method used for load carriage among sand miners (i.e. doko and namlo), our ability to compare the average load-carrying behaviors of sand miners to occupational load carrying in developed countries, such as the United States, is limited. Despite this, the loads carried by miners in our study were substantially higher than the 23 kg weight limit recommended by the National Institute for Occupational Safety and Health (NIOSH) for weights that can be lifted over an 8-hour period without increased risk of lower back injury (7,8). This comparison also does not consider sand miners’ deviations from “ideal” recommended lifting conditions (e.g., hand holds not available for lifting; twisting involved in lifting) that would further decrease the maximum acceptable weight limit recommended by NIOSH. To contextualize loads carried by female and male sand miners (average load weight: 66.3 kg and 87.4 kg, respectively), competitors in the 2000 World Masters Weightlifting Championships had an average combined total weight for the snatch lift and the clean and jerk lift of 43.94 kg among females and 54.22 among males (9). Although lifting methods used by weightlifters for barbell weights are quite different to lifting methods utilized by sand miners in Nepal, the average sand miner in our study population carried loads heavier than heavy weight lifting athletes for a reported average of 305 times per week, with an average duration of 2.7 minutes per load. Thus, the workloads of individuals in this study would be expected of only extremely fit workers. The limited variability in load-carrying exposures among the miners in our study population may have restricted our ability to identify associations between degrees of load carrying and musculoskeletal health. Alternatively, our findings may indicate the counterintuitive result that there was no adverse association between heavy load-carrying exposures and musculoskeletal health in our study population. To overcome this problem of inference, future studies examining the association between heavy load carrying and musculoskeletal health in this occupational group should aim to have larger sample sizes and include a comparable group of people not engaging in sand mining.

To explain the apparently inverse association between load weight and prevalence of any MSD (Tables VI and VII), we hypothesize that the extreme workload associated with carrying frequent, very heavy loads for long durations in our study population may be a driver of the healthy worker survivor bias (11). Essentially, sand miners experiencing the most severe musculoskeletal pain and MSDs would have since discontinued employment as sand miners and were subsequently unavailable to be recruited into our study. These findings are consistent with other research conducted in Nepal in which porters admitted to hospitals with head injuries over a two-year period had significantly reduced odds of radiologic cervical spondylosis, compared to non-porters also admitted with head-injuries (6). Although other studies assessing spinal problems in head-supported domestic load carriage have not identified protective associations between increasing load-carrying exposures and MSDs, comparison of our findings to other studies is limited by the extreme load weights and unique mechanisms of spinal loading associated with load carriage using dokos and namlos among our population of Nepalese men and women (3–6,12–15). Furthermore, studies examining domestic load carriage may be inherently less susceptible to the healthy worker survival effect, given that loads such as water and firewood for cooking are essential for normal living and, therefore, are carried regardless of musculoskeletal pain.

We identified higher prevalences of severe musculoskeletal pain and probable MSDs among females compared to males in our study population. The healthy worker survival effect may be less pronounced among women in our study, although our study size is small, severely limiting any such inference. Such differences between the sexes, if confirmed, might be because of the patriarchal societal structure in Nepal, possibly preventing women from leaving the occupation of sand mining despite experiencing severe musculoskeletal pain (16,17). Apart from load carriage as an income-generating activity, women in developing countries such as Nepal, are also likely to carry heavy loads throughout the year in order to keep their households supplied with necessities, such as water and fuel (3). Although a majority of women in our study did not report carrying loads such as water and firewood in the 7 days prior to the survey, women reported beginning to carry such loads as early as 4 years of age. The accumulation of domestic load-carrying exposures throughout women’s lives may have predisposed them to musculoskeletal injuries while working as sand miners.

Perceptions and reporting of pain also vary by sex, with evidence that women generally have a greater likelihood of reporting pain and poor health compared to men (18,19). Under- or over-reporting of existing musculoskeletal pain in our study could be subject to misclassification, since participants not reporting pain above a certain threshold were not evaluated by physical examination for the presence of MSDs. Under-reporting of pain by men would have biased our effect estimates towards or below the null. Our findings of generally high positive predictive values for moderate to severe pain in predicting true positive cases of probable MSDs in the neck and back (confirmed by physical examinations) suggests that self-reported pain may have reasonable enough diagnostic accuracy to be a surrogate for identifying MSDs in studies unable to conduct physical examinations or diagnostic testing due to funding or practical limitations. However, the high positive predictive values identified in our study may also be due in part to the high prevalence of MSDs in our study population (20). Additionally, we found a low diagnostic accuracy of self-reported pain in identifying MSDs in the knees, feet, and ankles. This may indicate the potential for outcome misclassification in studies that diagnose lower extremity musculoskeletal problems based only on self-reported pain, implying that physical examination of the lower extremities without supporting diagnostics (e.g., MRIs or x-rays) is insufficient to identify probable MSDs in these regions.

Musculoskeletal pain can exist secondary to rheumatological, metabolic and infectious causes, and MSD cases due to these causes may not correlate with occupational musculoskeletal load exposures (21). For example, potentially relevant to sand mining, occupational exposure to silica in sand has been associated with increased risk of rheumatoid arthritis (22). As our study did not attempt to diagnose or rule out the above-mentioned causes of musculoskeletal complaints, it is possible that some of our participants had MSDs due to such other causes. Our results may also be confounded by body mass index (BMI), given the higher prevalence of probable MSDs among miners with higher BMIs, although higher BMI may be a consequence of having an MSD. Our study did not use laboratory or radiological assessments to categorize MSD, and all exams were performed by one person, with no additional confirmation of physical exam findings. We expect any misclassification of MSD outcomes in our study to have occurred non-differentially with respect to load-carrying exposures and would have biased our results towards the null. Despite these potential limitations, the case criteria used were more specific than pain alone and more scalable for assessing associations between exposures and outcomes in populations where more expensive and/or invasive techniques are not practical.

The high prevalence of self-reported moderate to severe musculoskeletal pain and probable MSDs in our study population underscores the urgent need for future studies examining the adverse health effects of sand mining. According to World Health Organization Global Burden of Disease studies conducted in 2013 and 2015, musculoskeletal disorders, such as lower back pain and neck pain, represent the single largest cause of years lost to disability (YLD) globally (23,24). Notably, the burden of musculoskeletal disorders in terms of disability adjusted life years (DALYs) in developing countries is estimated to be 2.5 times that of developed countries (25). Although growing urbanization and increasing demand for raw building materials in developing countries seems likely to exacerbate this disparity, there has been little research on the extent of the problem among occupational groups like sand miners. Although we cannot easily compare our findings to other studies directly, given the paucity of epidemiologic data on sand-miners, our results are consistent with recent evidence from Tanzania suggesting increasing odds of musculoskeletal pain with increasing trip duration among women carrying domestic loads (e.g., water, wood, etc.) as part of their daily activities (10). While the magnitudes of our effect estimates suggest that load-carrying duration and frequency may contribute to the high prevalence of musculoskeletal health problems among sand miners, our estimates had wide confidence intervals that included the null, likely because of a small sample size. We provide, however, preliminary data that gives insight for design of larger, better-powered epidemiologic studies to more precisely explore causal relationships between load carrying by sand miners and MSDs, preferably using longitudinal designs. In addition to being larger, future studies building on our results should, if practical, employ physical examinations to diagnose probable MSDs among *all* individuals reporting pain, and should include supportive diagnostic testing when feasible.

## 5. CONCLUSION

We have conducted what we believe is the first epidemiologic study of Nepali sand miners, including physical examinations to confirm self-reported musculoskeletal pain. Despite our limited sample size, we have identified musculoskeletal disability problems affecting this virtually unstudied occupational group that is deserving of follow-up investigation in much larger studies. Although, because of the magnitude of the load weights carried, sand miners might be considered an extreme example, the occupational and general public health impacts of heavy load carrying in developing countries are barely, if at all, on the agendas of public health and occupational health authorities and funders. We hope that exploratory studies like ours will help to increase recognition of this widespread issue throughout the developing world. Especially needed is a source of funding for studies of heavy load carrying that can help to define the extent and magnitude of the problem, providing data to justify the development of preventive strategies.

## Data Availability

Data will be made available upon reasonable requests to the corresponding author.

## 7. ACKNOWLEDGEMENTS

We would like to thank the Bixby Center, as well as our many collaborators involved in the design and implementation of this study, particularly Jillian L. Kadota, Bina Gurung, Birendra Kunwar, and Dr. Dhirendra Prasad Bhatt. Most importantly, we would like to thank the sand miners who participated in this study, without whom this research would not have been possible.

